# Neuropsychological assessments for dementia research in the COVID-19 era: comparing remote and face-to-face testing

**DOI:** 10.1101/2022.04.28.22274370

**Authors:** Maï-Carmen Requena-Komuro, Jessica Jiang, Lucianne Dobson, Elia Benhamou, Lucy Russell, Rebecca L Bond, Emilie V Brotherhood, Caroline Greaves, Suzie Barker, Jonathan D Rohrer, Sebastian J Crutch, Jason D Warren, Chris JD Hardy

**Author notes:** joint lead authors (Maï-Carmen Requena-Komuro, Jessica Jiang). Correspondence to: Dr Chris J D Hardy, Dementia Research Centre, UCL Queen Square Institute of Neurology, 1^st^ Floor, 8-11 Queen Square, London WC1N 3AR, United Kingdom.

## Abstract

**Objectives:** We explored whether adapting traditional neuropsychological tests for online administration against the backdrop of COVID-19 was feasible for people with diverse forms of dementia and healthy older controls. We compared face-to-face and remote settings to ascertain whether remote administration affected performance.

**Design:** We used a longitudinal design for healthy older controls who completed face-to-face neuropsychological assessments between three and four years before taking part remotely. For patients, we used a cross-sectional design, contrasting a prospective remote cohort with a retrospective face-to-face cohort matched in age, education, and disease duration.

**Setting:** Remote assessments were performed using video-conferencing and online testing platforms, with participants using a personal computer or tablet and situated in a quiet room in their own home. Face-to-face assessments were carried out in dedicated testing rooms in our research centre.

**Participants:** The remote cohort comprised ten healthy older controls (also seen face-to-face 3-4 years previously) and 25 patients (n=8 Alzheimer’s disease (AD); n=3 behavioural variant frontotemporal dementia (bvFTD); n=4 semantic dementia (SD); n=5 progressive nonfluent aphasia (PNFA); n=5 logopenic aphasia (LPA)). The face-to-face patient cohort comprised 64 patients (n=25 AD; n=12 bvFTD; n=9 SD; n=12 PNFA; n=6 LPA).

**Primary and secondary outcome measures:** The outcome measures comprised the strength of evidence under a Bayesian analytic framework for differences in performances between face-to-face and remote testing environments on a general neuropsychological (primary outcomes) and neurolingustic battery (secondary outcomes).

**Results:** There was evidence to suggest comparable performance across testing environments for all participant groups, for a range of neuropsychological tasks across both batteries.

**Conclusions:** Our findings suggest that remote delivery of neuropsychological tests for dementia research is feasible.

**Strengths and limitations of this study:** *Methodological strengths of this study include:*

- Diverse patient cohorts representing rare dementias with specific communication difficulties
- Sampling of diverse and relevant neuropsychological domains
- Use of Bayesian statistics to quantify the strength of evidence for the putative null hypothesis (no effect between remote and face-to-face testing)

*Limitations include:*

- Relatively small cohort sizes
- Lack of direct head-to-head comparisons of test environment in the same patients

## 1. Introduction

The COVID-19 pandemic and associated social distancing and lockdown measures imposed a series of daunting challenges for conducting research with people with dementia. In the UK, three national lockdowns, imposed between March 2020 and February 2021 largely prevented face-to-face research. People with dementia are at increased risk for COVID-19 ^1^, and many participants understandably did not feel safe to travel for research purposes, particularly before widespread vaccination was implemented. Here, we describe our attempts to translate our traditional neuropsychology and neurolinguistics batteries (typically administered face-to-face) for remote administration.

Development and implementation of online cognitive assessments for dementia patients, particularly within communities who experience difficulties in accessing clinical care is not new ^2^. Telemedicine has been previously used successfully in the context of Alzheimer’s disease (AD) ^3 4^ and in people with rarer dementias such as primary progressive aphasia (PPA) ^5-7^ and behavioural variant frontotemporal dementia (bvFTD)^7^. However, with the COVID-19 pandemic, there has been a more pervasive shift towards the use of online methods to meet clinical, support and research needs ^8 9^.

A recent review by Hunter and colleagues ^10^ summarises 20 years of research comparing face-to-face and online administration of cognitive tests in healthy older adults (≥40 years old) and participants diagnosed with mild cognitive impairment (MCI), AD, or other types of dementia (often unspecified). The authors identified 12 studies that used videoconferencing methods. Overall, there was clear evidence to suggest that remote cognitive testing for people living with AD and other forms of dementia is feasible. In addition, there is evidence to suggest that online performance remains stable over time (with a maximum delay of three months between assessments), particularly for the domains of executive function, working memory, verbal episodic memory, and language. Minimal evidence was available for visuospatial tasks, and tests of single word and sentence comprehension.

Notwithstanding considerable progress in this area to date, further research into remote neuropsychological testing of patients with neurodegenerative diseases is required. There are three main issues that need addressing. First, it would be informative to understand more about the feasibility of remote testing in a non-controlled “home” environment (e.g., with potentially unstable internet connection and technological equipment not designed for research purposes). Second, it would be instructive to assess a range of patients with different forms of dementia, including PPA and bvFTD. Third, it is important to explore the feasibility of a range of neuropsychological tests measuring diverse cognitive functions remotely.

Based largely on the face-to-face protocol for general neuropsychological and neurolinguistic testing used at our research centre, we built a protocol for remote testing of patients diagnosed with typical AD, patients representing major variants of PPA (semantic dementia (SD), progressive nonfluent aphasia (PNFA), logopenic aphasia (LPA)), and bvFTD. Patients were tested directly from their homes via the widely used video conferencing software, Zoom (Zoom Video Communications Inc). We also recruited a small cohort of healthy older adults who had taken part in our face-to-face research at the Dementia Research Centre three to four years before the pandemic. Here, we compared the healthy controls’ performance on several neuropsychological and neurolinguistic tests between the two testing environments (face-to-face vs remote). We also compared the performance of patients tested remotely with a historical face-to-face cohort of patients chosen to represent the same syndromes and to match the remote cohort based on age, education, and symptom duration. We adopted a Bayesian approach that assesses the amount of evidence in favour of the null hypothesis (i.e., that there is no significant difference in performance on a given neuropsychological task between testing environments) relative to the alternative hypothesis (i.e., that there is a significant difference in performance on a given neuropsychological task between testing environments).

Following previous research ^10^, we did not predict major differences in terms of participants’ performances when tested face-to-face and remotely on most neuropsychological and neurolinguistics tests. However, we did consider the potential for poorer performance on tests of speech perception that were administered remotely, given additional difficulties associated with controlling the remote auditory environment.

## 2. Methods

### Participant recruitment and group matching

Recruitment for the study took place between February and August 2021. Potential patient participants were identified via the Specialist Cognitive Disorders Clinic at the National Hospital for Neurology and Neurosurgery, direct research referrals from external clinicians or via the Rare Dementia Support (www.raredementiasupport.org) network; healthy controls were recruited via our research participant database.

An initial telephone screen was conducted for each participant to establish they had access to the necessary equipment (tablet or desktop/laptop computer), a broadband internet connection, a quiet testing space to support the remote research assessment, and no preclusive hearing or visual impairments. We also performed the telephone version of the Mini-Mental State Examination (MMSE) with patients to assess their disease severity ^11 12^. A cut-off score of 12 on the T-MMSE (which corresponds to a converted MMSE score of 16) was used as an inclusion criterion ^11^. Of the potential participants who were approached about research, six (two healthy controls, one right temporal variant FTD, one PNFA, two LPA) declined for reasons related to remote delivery (e.g., not being comfortable with videoconferencing technology).

Twenty-five patients (eight with typical AD, three bvFTD, four SD, five PNFA, five LPA) were recruited for the remote study. For comparison purposes, a reference historical cohort comprising 64 patients (25 with AD, 12 bvFTD, nine SD, 12 PNFA, six LPA) who had undertaken a face-to-face research assessment at our Centre between 2013 and 2020 were selected, matching the cohort assessed remotely as closely as possible for syndromic composition, age, years of education and symptom duration. Henceforth, these are referred to as the ‘remote’ and ‘face-to-face’ patient cohorts, respectively. All patients fulfilled consensus diagnostic criteria for the relevant syndromic diagnosis ^13-15^ and all had clinically mild-to-moderate severity disease. Where available, brain MRI was consistent with the syndromic diagnosis, without evidence of significant cerebrovascular burden. Ten healthy older individuals with no history of neurological or psychiatric illness and who had been seen for face-to-face testing between three and four years previously also underwent remote assessments. Demographic and clinical details for all participants are summarised in Table 1.

**Table 1.** General demographic and clinical characteristics for all participant groups: comparison of remote and face-to-face cohort characteristics.

| Characteristic | CTL | AD |  | bvFTD |  | SD |  | PNFA |  | LPA |  |
| --- | --- | --- | --- | --- | --- | --- | --- | --- | --- | --- | --- |
|  | Remote | F2F | Remote | F2F | Remote | F2F | Remote | F2F | Remote | F2F | Remote |
| N | 10 | 25 | 8 | 12 | 3 | 9 | 4 | 12 | 5 | 6 | 5 |
| Gender (M/F) | 7/3 | 15/10 | 5/3 | 8/4 | 3/0 | 7/2 | 2/2 | 6/6 | 2/3 | 4/2 | 4/1 |
| Age (yrs) | 74.0<br>(4.1)* | 69.5<br>(4.1) | 69.8<br>(5.9) | 70.3<br>(2.9) | 72.0<br>(5.6) | 60.3<br>(5.2) | 58.3<br>(9.3) | 67.1<br>(5.2) | 68.2<br>(6.4) | 69.5<br>(3.6) | 71.6<br>(4.7) |
| Education (yrs) | 17.6<br>(0.7) | 14.8<br>(2.5) | 16.0<br>(3.7) | 14.9<br>(2.7) | 14.7<br>(3.1) | 16.0<br>(1.9) | 15.5<br>(3.1) | 15.3<br>(2.3) | 14.8<br>(2.8) | 16.0<br>(1.7) | 16.0<br>(3.3) |
| Handedness (R/L/A) | 9/1/0 | 23/1/1 | 8/0/0 | 11/1/0 | 3/0/0 | 9/0/0 | 3/1/0 | 10/2/0 | 5/0/0 | 5/1/0 | 5/0/0 |
| Symptom duration (yrs) | NA | 6.8<br>(2.3) | 7.3<br>(3.7) | 4.7<br>(2.1) | 3.7<br>(3.8) | 4.1<br>(1.9) | 3.5<br>(1.9) | 3.4<br>(1.7) | 3.2<br>(0.8) | 4.2<br>(1.9) | 3.8<br>(1.9) |

Mean (standard deviation) values are shown for each group. No significant differences between remote and face-to-face cohorts were found. \*on average healthy controls were 3.5 years younger when tested face-to-face; A, ambidextrous; AD, patient group with typical Alzheimer's disease; bvFTD, patient group with behavioural variant frontotemporal dementia; CTL, healthy control group; F, female; F2F, face-to-face; L, left; LPA, patient group with logopenic progressive aphasia; M, male; N, number of participants per group; NA, not applicable; PNFA, patient group with progressive non-fluent aphasia; R, right; SD, patient group with semantic dementia.

All participants gave informed consent for their involvement in the study. Ethical approval was granted by the University College London and National Hospital for Neurology and Neurosurgery Joint Research Ethics Committees in accordance with the Declaration of Helsinki.

### Testing procedure – face-to-face

Data for the reference historical cohort were collected under our face-to-face research assessment protocol, as delivered in experimental sessions at the Dementia Research Centre between 2013 and 2020. Under this protocol, all neuropsychological tests were administered in dedicated quiet testing rooms, with the participant sitting opposite the experimenter. Patients were predominantly tested on their own, unless the informant accompanying them to the study visit requested to be present and the participant agreed to this. In these cases, the informant was explicitly asked not to intervene during testing. No feedback was given on performance and no time limits were imposed (unless timing was intrinsic to the test). General neuropsychological and neurolinguistic test batteries (see Tables 2 and 3) were administered, following standard methods. The neurolinguistic battery was developed specifically to characterise the language profiles of people with PPA and therefore, was not administered to bvFTD or AD participants.

**Table 2.** General neuropsychological performance for all participant groups: comparison of remote and face-to-face test administration.

| Test | CTL |  | AD |  | bvFTD |  | SD |  | PNFA |  | LPA |  |
| --- | --- | --- | --- | --- | --- | --- | --- | --- | --- | --- | --- | --- |
|  | F2F | Remote | F2F | Remote | F2F | Remote | F2F | Remote | F2F | Remote | F2F | Remote |
| **No. tested** | 10 | 10 | 25 | 8 | 12 | 3 | 9 | 4 | 12 | 5 | 6 | 5 |
| *General intellect* |  |  |  |  |  |  |  |  |  |  |  |  |
| WASI Matrix (/32) | 26.9 (2.4) | 26.3 (3.5) | **14.5 (7.5)** | **13.7 (11.5)<sup>b</sup>** | 14.6 (9.5)<sup>a</sup> | 7.7 (2.5) | 25.8 (4.9) | 23.3 (3.4) | 18.6 (8.3)<sup>a</sup> | 24.0 (2.6) | 10.0 (9.2) | 17.6 (10.3) |
| *Episodic memory* |  |  |  |  |  |  |  |  |  |  |  |  |
| RMT faces (Short) (/25) | NA | 23.3 (2.5) | 18.9 (3.7)<sup>h</sup> | 15.3 (2.1) | NA | 20.3 (4.0) | NA | 19.3 (3.7) | NA | 22.4 (3.7) | NA | 21.6 (3.8) |
| RMT faces (/50) | 42.7 (4.7) | NA | 30.44 (6.56)<sup>g</sup> | NA | 28.56 (6.29)<sup>c</sup> | NA | 31.5 (4.47)<sup>a</sup> | NA | 40.3 (5.79)<sup>b</sup> | NA | 33.17 (8.93) | NA |
| *Working memory* |  |  |  |  |  |  |  |  |  |  |  |  |
| DS (Forward) (/12) | 9.8 (2.0) | 9.2 (2.0) | **6.5 (2.1)<sup>a</sup>** | **6.6 (2.3)** | 7.8 (2.9) | 6.3 (0.6) | 9.4 (2.2) | 7.3 (0.5) | 5.1 (2.6)<sup>b</sup> | 6.0 (4.4) | 3.0 (3.1)<sup>a</sup> | 5.0 (2.5) |
| *Language* |  |  |  |  |  |  |  |  |  |  |  |  |
| BPVS (/150) | **149.0 (1.1)** | **148.9 (1.1)** | **135.1 (24.0)<sup>a</sup>** | **133.9 (21.4)** | 118.7 (28.9)<sup>a</sup> | 148.0 (1.0) | 61.9 (44.1) | 98.0 (46.3) | 136.3 (30.4) | 142.8 (8.4) | 120.2 (39.6) | 145.2 (4.4) |
| GNT (/30) | 27.3 (2.1) | 26.8 (1.8) | **13.7 (8.5)<sup>a</sup>** | **12.4 (6.3)** | 11.5 (11.0)<sup>a</sup> | 20.0 (4.4) | 0.2 (0.4) | 0.5 (1.0) | 15.8 (6.8) | 20.2 (7.6) | 6.2 (7.7) | 12.4 (7.0) |
| NART (/50) | 45.2 (3.4) | 44.6 (3.0) | 28.3 (11.9)<sup>a</sup> | 37.1 (4.6) | 30.7 (13.1) | 40.0 (2.0) | 16.1 (10.2) | 16.3 (13.6) | 28.3 (15.0)<sup>f</sup> | 30.8 (14.9) | 20.4 (18.5)<sup>a</sup> | 29.8 (16.9) |
| *Arithmetic* |  |  |  |  |  |  |  |  |  |  |  |  |
| GDA Total (/24) | **16.6 (6.4)** | **16.1 (4.0)** | **4.9 (5.0)<sup>e</sup>** | **4.8 (5.4)** | 8.2 (7.4)<sup>a</sup> | 8.3 (7.2) | 13.3 (5.5) | 7.3 (4.0) | 5 (3.6)<sup>d</sup> | 6.4 (7.2) | 1.7 (1.5) | 7 (5.8) |
| *Visuospatial* |  |  |  |  |  |  |  |  |  |  |  |  |
| VOSP OD (/20) | *19.5 (0.7)* | *18.4 (1.2)* | *16.1 (2.6)<sup>a</sup>* | *13.0 (3.0)* | 14.0 (4.2) | 17.0 (1.0) | 15.3 (6.3)<sup>a</sup> | 14.5 (5.9) | 16.8 (3.6)<sup>b</sup> | 17.6 (2.1) | 15.8 (3.1) | 15.8 (2.7) |
| *Executive* |  |  |  |  |  |  |  |  |  |  |  |  |
| DS (Reverse) (/12) | **8.0 (2.6)** | **8.2 (2.2)** | 4.6 (1.9)<sup>c</sup> | 4.0 (1.9) | 5.0 (3.1)<sup>a</sup> | 4.0 (0.0) | 6.9 (2.7) | 5.5 (2.7) | 4.4 (1.7)<sup>d</sup> | 5.2 (4.2) | 2.4 (1.1)<sup>a</sup> | 4.8 (1.8) |
| Letter fluency (60s, “F”) | **19.7 (6.0)** | **20.8 (4.2)** | 10.3 (5.2) | 11.9 (7.4) | 6.8 (3.9)<sup>a</sup> | 7.3 (2.3) | 7.0 (3.6)<sup>b</sup> | 10.8 (7.0) | 4.8 (5.8)<sup>d</sup> | 14.0 (8.2)<sup>a</sup> | 2.6 (3.8)<sup>a</sup> | *11.3 (3.9)* |
| Category fluency (60s, “Animals”) | **25.0 (6.6)** | **24.7 (5.4)** | **10.0 (5.6)** | **10.4 (6.7)** | 8.0 (6.1) | 10.3 (6.0) | 6.1 (5.3)<sup>a</sup> | 9.8 (6.2) | 8.6 (5.8)<sup>d</sup> | 18.6 (11.5) | 5.7 (5.8) | 11.6 (8.1) |

**Table 3.**
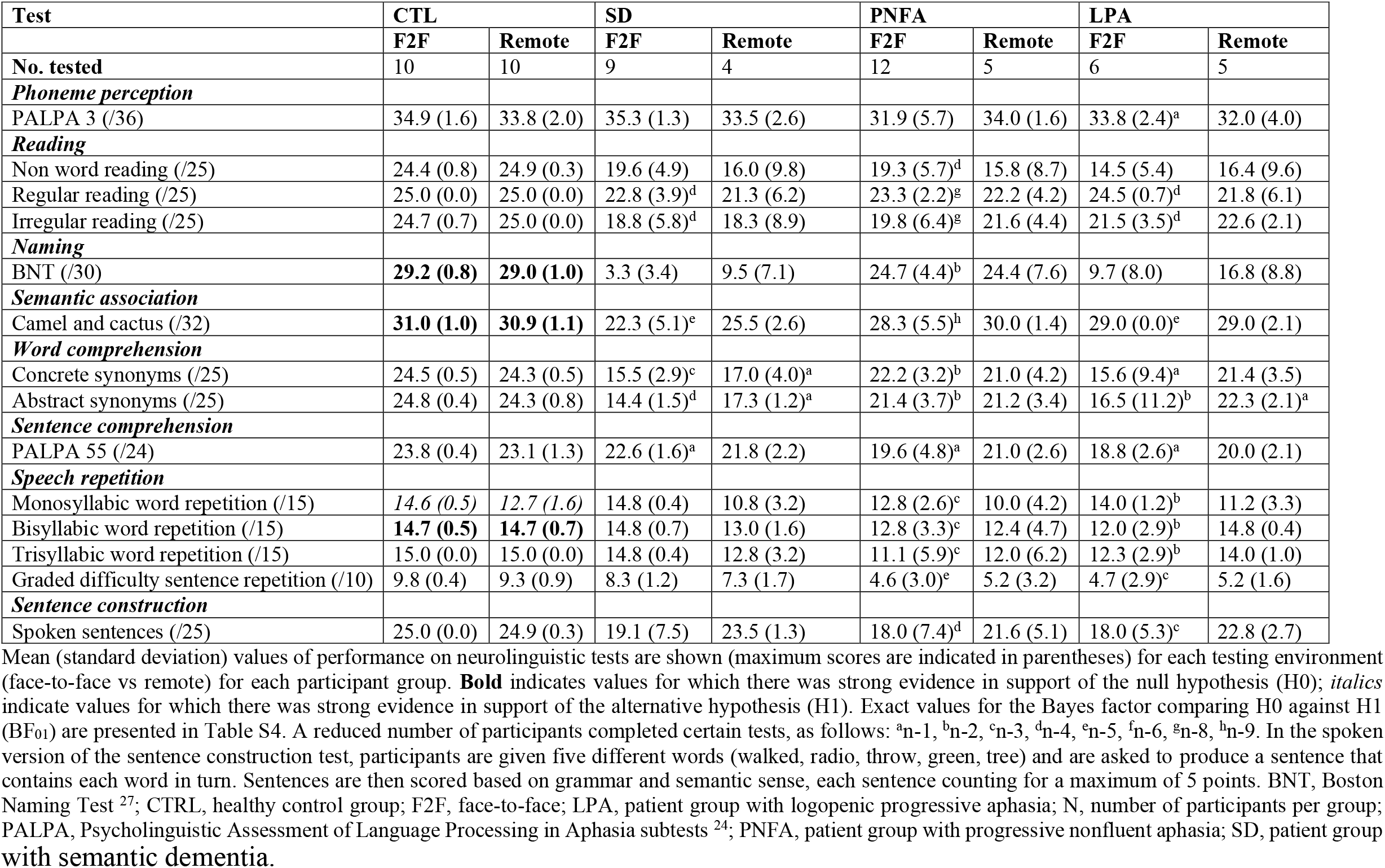
Neurolinguistic performance for all participant groups: comparison of face-to-face and remote test administration.

| Test | CTL |  | SD |  | PNFA |  | LPA |  |
| --- | --- | --- | --- | --- | --- | --- | --- | --- |
|  | F2F | Remote | F2F | Remote | F2F | Remote | F2F | Remote |
| **No. tested** | 10 | 10 | 9 | 4 | 12 | 5 | 6 | 5 |
| ***Phoneme perception*** |  |  |  |  |  |  |  |  |
| PALPA 3 (/36) | 34.9 (1.6) | 33.8 (2.0) | 35.3 (1.3) | 33.5 (2.6) | 31.9 (5.7) | 34.0 (1.6) | 33.8 (2.4)<sup>a</sup> | 32.0 (4.0) |
| ***Reading*** |  |  |  |  |  |  |  |  |
| Non word reading (/25) | 24.4 (0.8) | 24.9 (0.3) | 19.6 (4.9) | 16.0 (9.8) | 19.3 (5.7)<sup>d</sup> | 15.8 (8.7) | 14.5 (5.4) | 16.4 (9.6) |
| Regular reading (/25) | 25.0 (0.0) | 25.0 (0.0) | 22.8 (3.9)<sup>d</sup> | 21.3 (6.2) | 23.3 (2.2)<sup>g</sup> | 22.2 (4.2) | 24.5 (0.7)<sup>d</sup> | 21.8 (6.1) |
| Irregular reading (/25) | 24.7 (0.7) | 25.0 (0.0) | 18.8 (5.8)<sup>d</sup> | 18.3 (8.9) | 19.8 (6.4)<sup>g</sup> | 21.6 (4.4) | 21.5 (3.5)<sup>d</sup> | 22.6 (2.1) |
| ***Naming*** |  |  |  |  |  |  |  |  |
| BNT (/30) | **29.2 (0.8)** | **29.0 (1.0)** | 3.3 (3.4) | 9.5 (7.1) | 24.7 (4.4)<sup>b</sup> | 24.4 (7.6) | 9.7 (8.0) | 16.8 (8.8) |
| ***Semantic association*** |  |  |  |  |  |  |  |  |
| Camel and cactus (/32) | **31.0 (1.0)** | **30.9 (1.1)** | 22.3 (5.1)<sup>e</sup> | 25.5 (2.6) | 28.3 (5.5)<sup>h</sup> | 30.0 (1.4) | 29.0 (0.0)<sup>e</sup> | 29.0 (2.1) |
| ***Word comprehension*** |  |  |  |  |  |  |  |  |
| Concrete synonyms (/25) | 24.5 (0.5) | 24.3 (0.5) | 15.5 (2.9)<sup>c</sup> | 17.0 (4.0)<sup>a</sup> | 22.2 (3.2)<sup>b</sup> | 21.0 (4.2) | 15.6 (9.4)<sup>a</sup> | 21.4 (3.5) |
| Abstract synonyms (/25) | 24.8 (0.4) | 24.3 (0.8) | 14.4 (1.5)<sup>d</sup> | 17.3 (1.2)<sup>a</sup> | 21.4 (3.7)<sup>b</sup> | 21.2 (3.4) | 16.5 (11.2)<sup>b</sup> | 22.3 (2.1)<sup>a</sup> |
| ***Sentence comprehension*** |  |  |  |  |  |  |  |  |
| PALPA 55 (/24) | 23.8 (0.4) | 23.1 (1.3) | 22.6 (1.6)<sup>a</sup> | 21.8 (2.2) | 19.6 (4.8)<sup>a</sup> | 21.0 (2.6) | 18.8 (2.6)<sup>a</sup> | 20.0 (2.1) |
| ***Speech repetition*** |  |  |  |  |  |  |  |  |
| Monosyllabic word repetition (/15) | *14.6 (0.5)* | *12.7 (1.6)* | 14.8 (0.4) | 10.8 (3.2) | 12.8 (2.6)<sup>c</sup> | 10.0 (4.2) | 14.0 (1.2)<sup>b</sup> | 11.2 (3.3) |
| Bisyllabic word repetition (/15) | **14.7 (0.5)** | **14.7 (0.7)** | 14.8 (0.7) | 13.0 (1.6) | 12.8 (3.3)<sup>c</sup> | 12.4 (4.7) | 12.0 (2.9)<sup>b</sup> | 14.8 (0.4) |
| Trisyllabic word repetition (/15) | 15.0 (0.0) | 15.0 (0.0) | 14.8 (0.4) | 12.8 (3.2) | 11.1 (5.9)<sup>c</sup> | 12.0 (6.2) | 12.3 (2.9)<sup>b</sup> | 14.0 (1.0) |
| Graded difficulty sentence repetition (/10) | 9.8 (0.4) | 9.3 (0.9) | 8.3 (1.2) | 7.3 (1.7) | 4.6 (3.0)<sup>e</sup> | 5.2 (3.2) | 4.7 (2.9)<sup>c</sup> | 5.2 (1.6) |
| ***Sentence construction*** |  |  |  |  |  |  |  |  |
| Spoken sentences (/25) | 25.0 (0.0) | 24.9 (0.3) | 19.1 (7.5) | 23.5 (1.3) | 18.0 (7.4)<sup>d</sup> | 21.6 (5.1) | 18.0 (5.3)<sup>c</sup> | 22.8 (2.7) |

with semantic dementia.

### Modifying the face-to-face neuropsychological battery for remote delivery

We reviewed the general neuropsychological and neurolinguistic batteries that had been used historically at our Centre for face-to-face administration, in order to identify tests that could be feasibly delivered remotely online while preserving the overall structure of the batteries and sampling across cognitive domains as far as possible. In selecting tests, an important consideration was to sample cognitive domains representatively so that we could establish phenotypic profiles of deficits in the target neurodegenerative syndromes (see Table S1). Where a task required visual stimulus presentation, a high-quality copy of the stimuli was made. Images were then imported into Microsoft Powerpoint for subsequent presentation to the participant via screen share.

Tests that were retained for remote testing (i.e., tests administered to both the remote and face-to-face patient cohorts) are itemised in Tables 2 and 3.The general neuropsychological battery comprised tests of general intellect (Wechsler Adult Intelligence Scale (WASI) Matrix Reasoning ^16^), episodic memory (Recognition Memory Test (RMT) for Faces ^17^), working memory (Digit span forwards ^18^), language (the graded naming test (GNT ^19^), British Picture Vocabulary Scale (BPVS ^20^) and National Adult Reading Test (NART) ^21^), arithmetic (Graded Difficulty Arithmetic (GDA)), visual perception (Visual Object and Space Perception (VOSP) Object Decision task ^22^) and executive function (Digit span reverse ^18^, letter (‘F’) and category fluency (‘animal’) tasks ^23^).

The neurolinguistic battery comprised tests of phoneme perception (a shortened version of the Psycholinguistic Assessments of Language Processing in Aphasia (PALPA) Test 3 – minimal pair discrimination ^24^), reading (graded non-word reading ^25^, and graded tests of regular word and irregular word reading adapted from ^26^), confrontation naming (a subset of items from the Boston Naming Test (BNT) ^27^), semantic association (modified Camel and Cactus test ^28^), single word comprehension (concrete and abstract synonyms ^29^), sentence comprehension (a shortened version of the PALPA Subtest 55 ^24^), speech repetition (tests of monosyllabic, bisyllabic, and trisyllabic single word repetition ^30^, graded difficulty sentence repetition using a subset of items from the Sentence Repetition test ^31^ and sentence construction based on ^32^. We opted to include two naming tasks as the GNT is part of the core neuropsychological battery at our Centre and therefore administered to patients with all diagnoses. However, as patients with SD often score at floor on this task ^33^, we also include a shortened version of the BNT in our neurolinguistic battery that is administered to patients with PPA as SD patients are more likely to be above floor on this task ^34^.

Where applicable, we sought permission from the test publishers to adapt tests for remote administration.

### Testing procedure – remote

An initial session was conducted via Zoom to accustom participants to the remote testing format, check the screen and sound sharing options on Zoom, and that the quality of their internet connection was acceptable.

Participants were permitted to use their preferred device (tablet, laptop, or desktop computer). To ensure screen visibility, we did not accept the use of smartphones. Most participants (90%) listened via speakers; six participants used headphones, and device volume was set to a comfortable level by each participant or their caregiver. Remote assessments were scheduled to ensure that testing could be completed in a quiet environment with minimal distractions. Where a task required visual stimulus presentation, this was done by screen sharing the Microsoft Powerpoint presentation containing the scanned stimuli for that task in full screen mode. Each patient’s primary caregiver was asked to be available during each research session in case of any problem occurs with using the equipment; in practice, no major technological issues arose.

To provide a check on basic audibility in the remote testing environment, before each remote testing session, participants first listened to a set of 10 sentences from the Bamford-Kowal-Bench (BKB) list ^35^. These sentences have previously been validated in hearing-impaired children. The spoken sentences were delivered online using an online experiment builder, Labvanced ^36^. In each trial, a spoken sentence was played to the participant via screen and sound share on Zoom and the participant was encouraged to select the last word in the sentence they had just heard from three possible options presented visually via screen share (see Figure S1). A perfect score on the final three items was required for the participant to proceed to the remote testing session proper (this allowed each participant and/or caregiver to manually adjust the volume to a comfortable level during the first seven sentences). Most participants (95%) performed at ceiling across all ten items, and no participant made an error on any of the final three items, meaning that none was rejected based on their BKB performance (see Table S2). The order of sentences was fixed across participants.

The remote neuropsychological and neurolinguistic batteries each took around an hour to administer. To minimise fatigue ^37^, batteries were delivered in separate testing sessions typically within a week (and never more than two weeks apart).

At the end of each testing session, each participant was debriefed by the experimenter. This provided them with the opportunity to raise any technical issues and give their impressions of the remote testing session. No technical difficulties were reported, and all participants said they had felt comfortable with remote testing.

### Statistical analysis

All statistical analyses were performed in JASP (version 0.16).

The remote and face-to-face patient cohorts were compared on demographic characteristics using independent samples t-tests and Wilcoxon rank-sum tests. Healthy controls’ scores in remote and face-to-face testing environments were compared using paired samples t-tests or (where the assumption of normality was not met) Wilcoxon signed rank tests. To reduce Type I error, no corrections for multiple comparisons were applied.

We did not perform between group comparisons of neuropsychological and neurolinguistic performance as these syndromic profiles of the neuropsychological and neurolinguistic tests have been reviewed and published previously ^38 39^.

Our null hypothesis was that there would be no effect of testing environment on neuropsychological performance – i.e., no differences in performance between remote and face-to-face assessment settings – for any participant group. To critically assess the magnitude of evidence in favour of this null hypothesis versus the alternative hypothesis (i.e., that there was in fact an effect of testing environment) particularly in light of the relatively small patient cohorts here, we employed a Bayesian approach ^40^. Bayesian independent samples t-tests (and non-parametric equivalents where assumptions of the general linear model were violated) were performed for each general neuropsychological and neurolinguistic test in each patient group separately. As numbers in some groups were quite small, we also conducted analyses for a combined patient cohort in both environments. Healthy control performance was compared using Bayesian paired samples t-tests (or appropriate non-parametric equivalent). A Bayes factor, which is the ratio of evidence supporting the null hypothesis over the alternative hypothesis (hereafter BF_01_) was calculated for each comparison using JASP. A BF_01_ value > 3 indicates strong evidence in favour of the null hypothesis while a value < 0.33 supports the alternative hypothesis; BF_01_ values between 0.33 and 3 are classified as ‘anecdotal’ evidence, comparable to non-significant differences in inferential statistics ^41 42^. Bayes factor values are presented in Tables S3 and S4.

### Patient and public involvement

In August and September 2020, we contacted 527 people (comprising healthy control participants and people with a diagnosis of a dementia) who had previously taken part in our face-to-face research programmes in the Dementia Research Centre, UCL, or who had expressed an interest in doing so in the future. They were asked, “Would you consider participating in research remotely (telephone/ online)?” Of the 163 people who answered the question, 145 (89%) indicated that they would be happy to take part in remote research. Based on this feedback, we submitted an amendment to our existing research ethics that was approved in October 2020. Following this, we conducted a pilot remote testing session with an older healthy control individual who was also a carer for a family member living with dementia. Their feedback was instrumental in developing and improving our remote testing procedure.

Results from this work will be disseminated to members of the support groups that we run with Rare Dementia Support (www.raredementiasupport.org) through online presentations at webinars and research summaries in newsletters.

## 3. Results

### General characteristics of participant groups

There were no significant differences in age, years of education or symptom duration between the face-to-face and remote testing patient cohorts (Table 1).

Below we highlight comparisons where there was strong evidence in support of either the null (i.e. no difference between remote and face-to-face performance) or alternative (i.e. difference between remote and face-to-face performance) hypothesis. Comparisons are shown in full in Tables S3 and S4.

### General neuropsychological assessment

Overall, there was little evidence for a significant effect of assessment environment on general neuropsychological test performance in any participant group.

Healthy individuals scored equally well on the digit span reverse, the British Picture Vocabulary Scale (BPVS), the Graded Difficulty Arithmetic (GDA), and on both letter and category fluency tests (all BF_01_ > 3 indicating strong evidence in favour of the null hypothesis). However, they performed less well on the Visual Object and Spatial Perception Object Decision task (VOSP) (BF_01_ = 0.0404, indicating strong evidence in favour of the alternative hypothesis) in remote testing than in face-to-face testing, though absolute performance differences were relatively small (remote mean = 18.4; face-to-face mean = 19.5; Tables 2 and S3, Figures 1 and 3).

**Figure 1.**
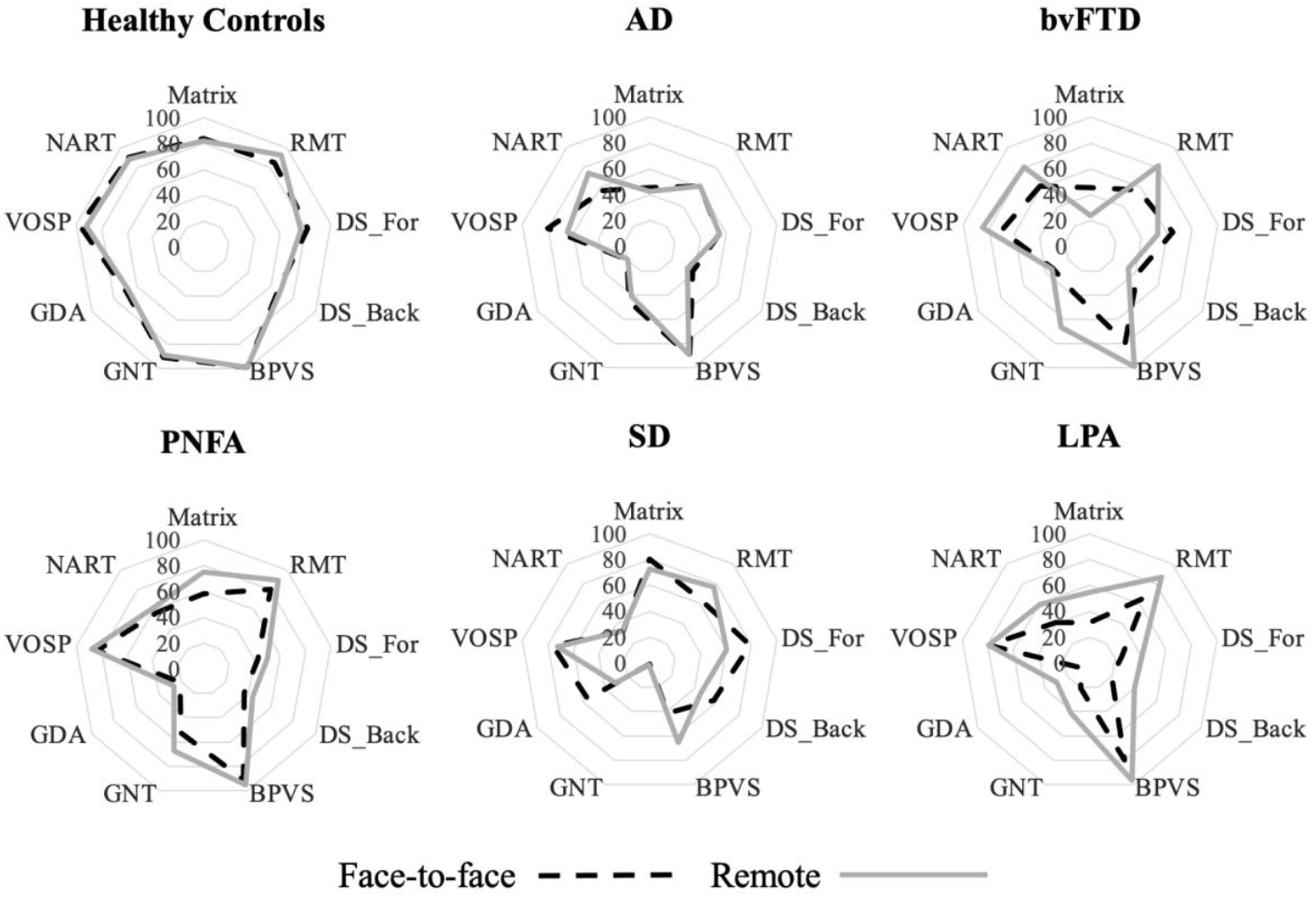
Radar plots of general neuropsychology battery performance, by participant group and testing environment. Average percentage correct score (plotted on concentric lines) was calculated for each participant group for each test in the neuropsychology battery, across each testing environment. Scores for the fluency tasks were not included here as responses on these tasks cannot be evaluated as correct/incorrect. AD, patient group with typical Alzheimer’s disease; BPVS, British Picture Vocabulary Scale; bvFTD, patient group with behavioural variant frontotemporal dementia; DS_For/Back, Digit Span Forwards/Backwards; GDA, Graded Difficulty Arithmetic test; GNT, Graded Naming Test; LPA, patient group with logopenic progressive aphasia; Matrix, WASI Matrix Reasoning; NART, National Adult Reading Test; PNFA, patient group with progressive nonfluent aphasia; RMT, Recognition Memory Test; SD, patient group with semantic dementia; VOSP, Visual Object Space Perception battery.

For the comparisons of the combined remote vs combined face-to-face patient cohorts, there was strong evidence supporting the null hypothesis for all neuropsychological tests (all BF_01_ > 3), except for the NART and both letter and category fluency tests, where evidence in support of the null hypothesis was anecdotal (Tables 2 and S3).

In respect of the individual patient groups (Tables 2 and S3, Figure 1), there was strong evidence to suggest that the AD group tested remotely performed similarly to the AD group seen face-to-face on WASI matrix reasoning, digit span forward, graded naming test (GNT), BPVS, GDA, and category fluency test (all BF_01_ > 3). However, the remote AD cohort performed less well on the VOSP (mean = 13.0; BF_01_ = 0.171, strong evidence) compared to the face-to-face cohort (mean = 16.1). Conversely, LPA patients who completed the letter fluency test remotely (mean words = 11.3) performed better than those who completed the same task face-to-face (mean = 2.6; BF01 = 0.188, strong evidence).

No other comparisons yielded strong evidence in support of either hypothesis (Table S3).

### Neurolinguistic assessment

Overall, there was little evidence for a significant effect of assessment environment on neurolinguistic test performance in any participant group.

Healthy individuals scored equally well on the Boston Naming Test (BNT), the camel and cactus test, and the bisyllabic single word repetition test (all BF_01_ > 3, indicating strong evidence in favour of the null hypothesis). However, they performed less well on the monosyllabic word repetition test (BF01 = 0.0487, strong evidence in favour of the alternative hypothesis) in remote testing than in face-to-face testing, though the absolute performance difference was quite small (remote mean = 12.7; face-to-face mean = 14.6; Tables 3 and S4, Figure 2).

**Figure 2.**
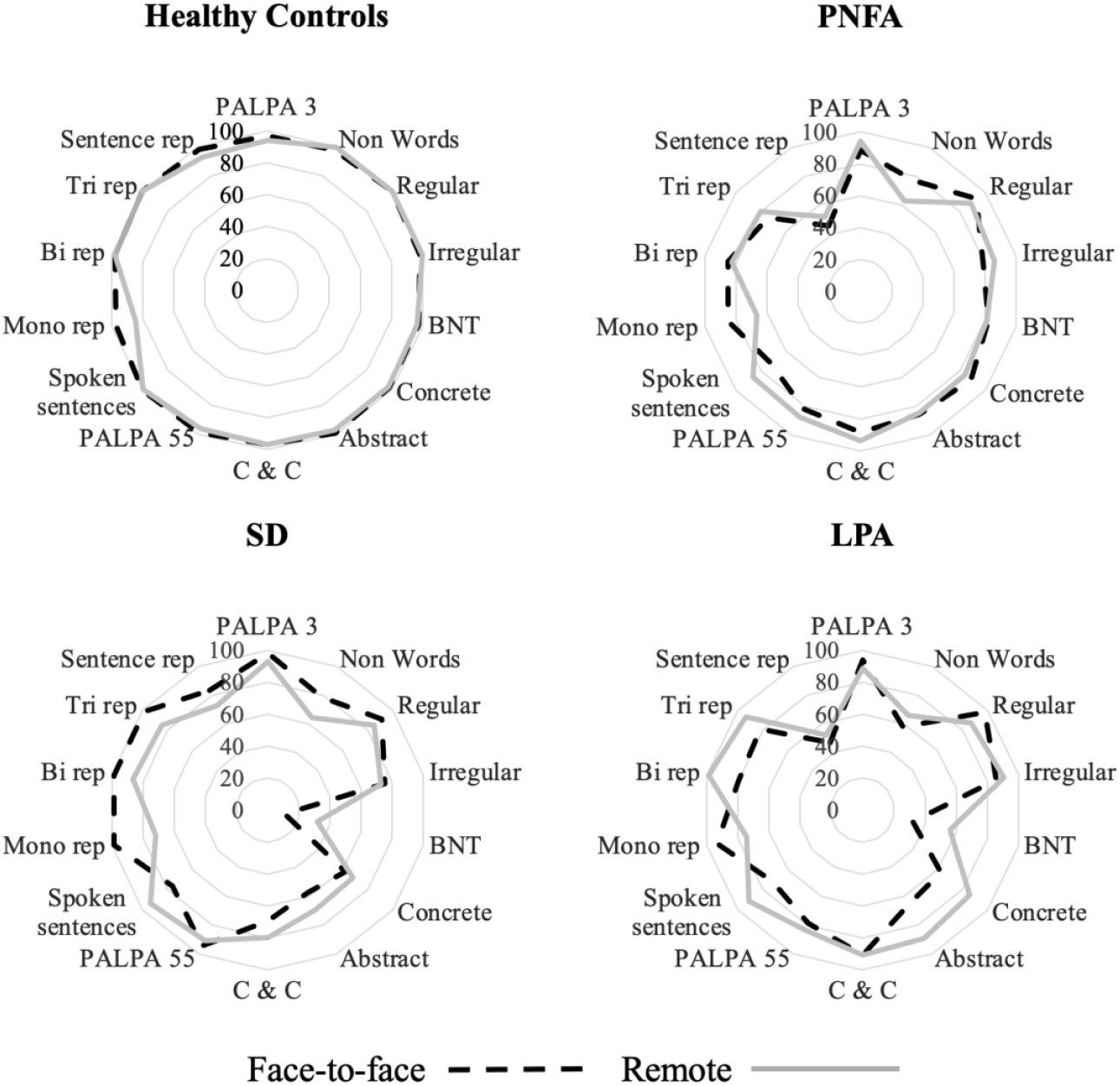
Radar plots of performance on neurolinguistic battery performance, by participant group and testing environment. Average percentage score (plotted on concentric lines) was calculated for each participant group for each test in the neurolinguistic battery, across each testing environment. Abstract, abstract synonyms test; bi rep, bisyllabic single word repetition; BNT, Boston Naming Test; C & C, camel and cactus test; concrete, concrete synonyms test; irregular, irregular word reading test; LPA, patient group with logopenic progressive aphasia; mono rep, monosyllabic single word repetition test; non word, non-word reading test; PALPA, Psycholinguistic Assessment of Language Processing in Aphasia subtests; PNFA, patient group with progressive nonfluent aphasia; regular, regular word reading test; sentence rep, graded difficulty sentence repetition test; SD, patient group with semantic dementia; spoken sentences, spoken sentences test; tri rep, trisyllabic single word repetition test.

**Figure 3.**
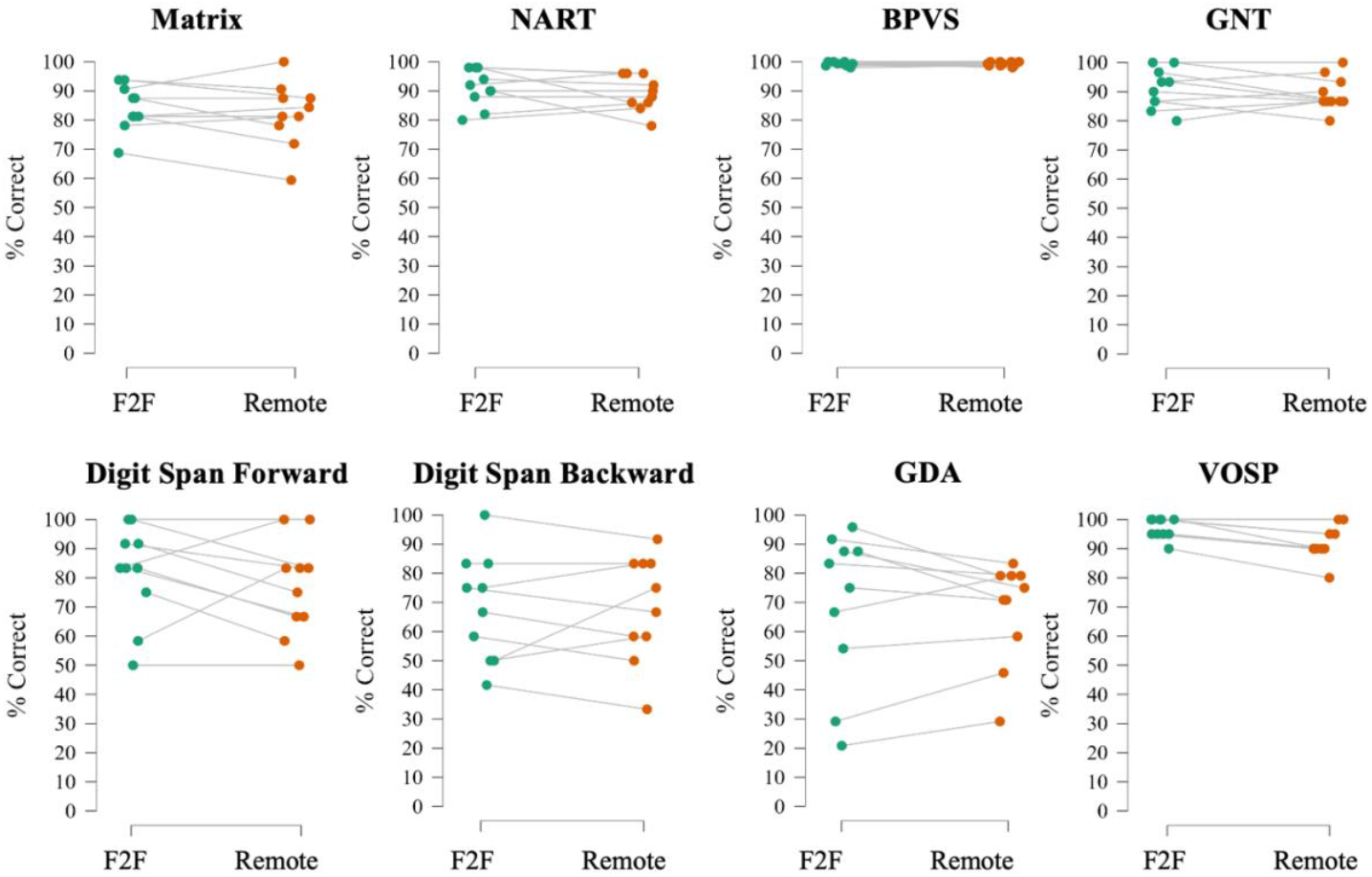
Performance profiles of healthy control participants on tasks in general neuropsychological battery. Line plots showing performance profiles of individual healthy control participants on tasks in the general neuropsychological battery. BPVS, British Picture Vocabulary Scale; GDA, Graded Difficulty Arithmetic test; GNT, Graded Naming Test; Matrix, WASI Matrix Reasoning; NART, National Adult Reading Test; VOSP, Visual Object Space Perception Object Decision task.

**Figure 4.**
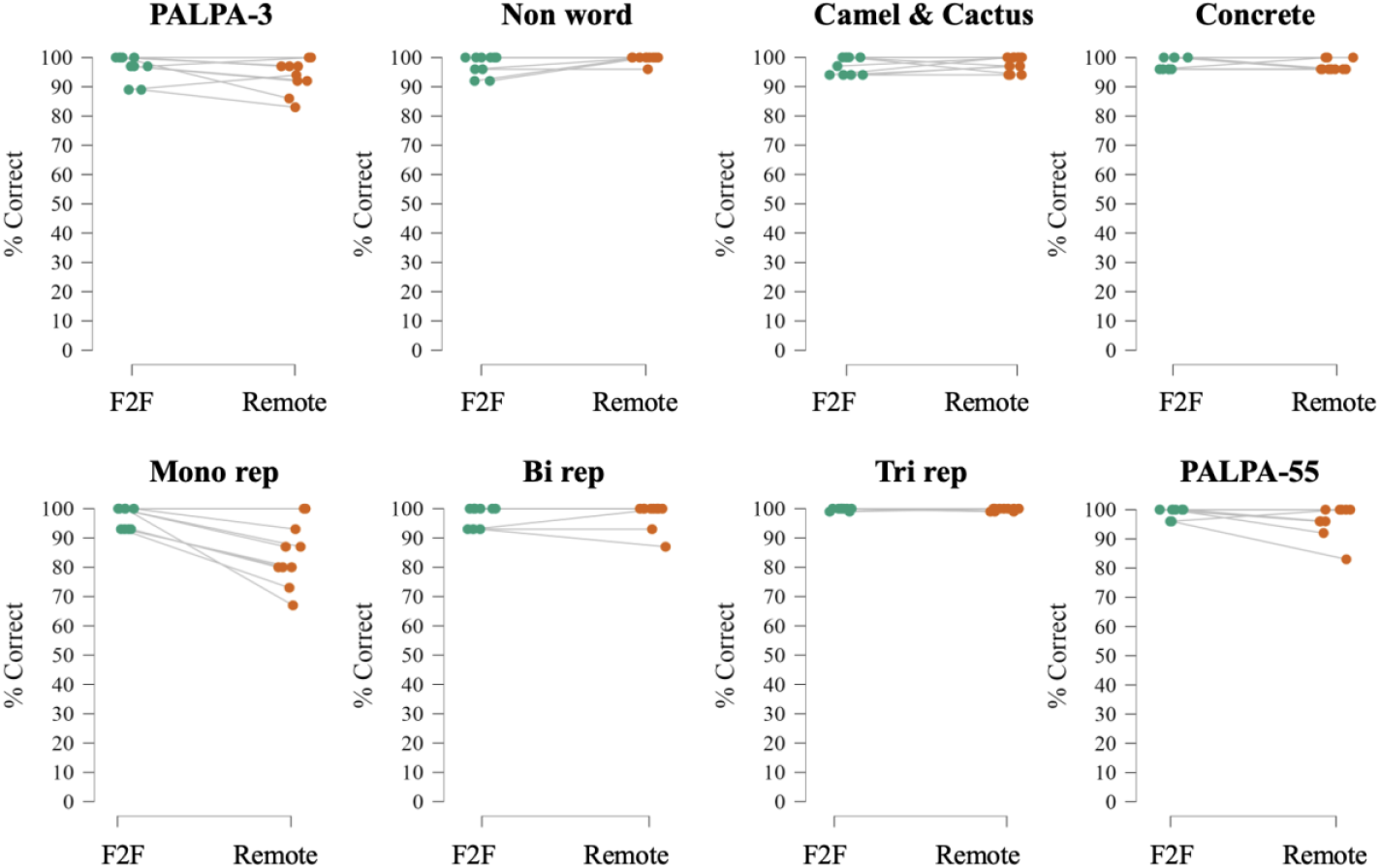
Performance profiles of healthy control participants on tasks in the neurolinguistic battery. Line plots indicating percentage scores for each healthy control on representative tests from the neurolinguistic battery administered face-to-face and remotely. Scores on the trisyllabic single word repetition task were jittered slightly on the x-axis to allow for plotting as participants were uniformly at ceiling in both environments. Bi rep, bisyllabic single word repetition; BNT, Boston Naming Test; Concrete, concrete synonyms test; F2F, face-to-face; Mono rep, monosyllabic single word repetition test; Non word, non-word reading test; PALPA, Psycholinguistic Assessment of Language Processing in Aphasia; Tri rep, trisyllabic single word repetition.

The comparisons between combined patient cohorts for remote vs face-to-face testing showed strong evidence supporting the null hypothesis for non-word reading, concrete synonyms, the PALPA-55, and bisyllabic and trisyllabic single word repetition tests (all BF_01_ values > 3). There was anecdotal evidence supporting the null hypothesis on all other neurolinguistic tests (all BF_01_ values higher between 1 and 3).

Individual patient group comparisons across environments did not yield strong evidence in support of either hypothesis.

## 4. Discussion

The present findings suggest that administration of neuropsychological tasks remotely over the internet with healthy older adults and people with a diverse range of dementia phenotypes is broadly feasible.

Our Bayesian analytical approach demonstrated that there was anecdotal or strong evidence suggesting comparable performance across testing environments of healthy participants and AD patients on a range of general neuropsychological and neurolinguistic tests, specifically those targeting working memory (digit span forward), executive functioning (digit span reverse, letter and category fluency tests, WASI matrix reasoning), arithmetic skills (GDA), and general semantic knowledge (BNT, BPVS, GNT). These results corroborate previous reports of preserved neuropsychological performance on executive function, working memory, and language tests across testing environments in both healthy individuals and AD patients ^10^. Our findings also corroborated previous work suggesting that remote assessments are viable for people with PPA ^43^.

Healthy controls and AD participants both performed significantly worse on the remote version of the VOSP object decision task, in which participants are presented with four silhouettes and asked to select the drawing of a real object; the three distractor silhouettes are based on nonsense shapes. The typical amnestic AD phenotype can include prominent visuospatial impairments ^44 45^ and it is feasible that a reduction in stimulus quality may have stressed cortical apperceptive mechanisms still further, akin to a dynamic ‘stress test’ of degraded input processing ^34 46 47^. However, it is worth noting that there was no such discrepancy across the AD cohorts for other tasks involving visual administration (e.g., WASI Matrix). For the healthy controls, the absolute performance difference across environments was relatively small (mean reduction of 1.1 points). It is also possible that this reduction at least in part reflected normal healthy ageing, as the healthy control cohort was tested on the remote battery between three to four years after their face-to-face assessment.

Healthy control participants also performed significantly worse on the monosyllabic single word repetition task when delivered remotely. The videoconferencing software may have degraded the fidelity of the raw speech signal ^48^, essentially resulting in a harder task than when administered face-to-face. This is potentially consistent with the controls’ preserved performance on the bisyllabic single word repetition test where top-down information can be used to complement bottom-up auditory information partially degraded by the videoconferencing software ^49^. An alternative (or complementary) explanation could again be age-related changes, here affecting hearing function (presbycusis) ^50-52^.

The finding of significantly better performance on the verbal (letter) fluency task in the LPA cohort tested remotely compared to the cohort tested face-to-face is surprising. The obvious explanation is that the remote cohort was overall less impaired than the patients seen face-to-face; although efforts were made to match the two cohorts for disease severity and other potentially relevant factors. An alternative explanation could be that participants found the remote setting less anxiety-provoking than face-to-face testing in an unfamiliar environment. Patients with LPA may be relatively susceptible to anxiety as a factor modulating cognitive performance ^53 54^. Additionally, as word retrieval is an intrinsically dynamic process that is likely to be facilitated by the availability of ‘prompts’ ^55^, patients may have benefitted from cueing of word retrieval by their more familiar home environments.

The way the neuropsychology and neurolinguistic testing protocol was adapted for remote delivery may have favoured null differences. The testing sessions were shorter and spread out within a week, which may have helped counteract the effect of anxiety related to the unfamiliarity of the remote testing setting, as well as potential ‘Zoom’ fatigue ^37^. The increased flexibility of scheduling compared to face-to-face testing in addition to the absence of potentially stressors associated with a face-to-face research visit (e.g., travelling, being in a unfamiliar environment) may have led to participants feeling more relaxed when taking part remotely vs face-to-face: future research should explore this. Furthermore, certain tests selected for remote delivery may have been intrinsically less susceptible to changes in testing protocol (e.g., BPVS), whereas we deliberately excluded tests that we considered would not be practical or suboptimal for remote delivery (e.g., WASI Block design, Baxter spelling test, Trails). Anecdotally, participants reported satisfaction with the remote testing protocol.

The current study presents several limitations which should inform future work. First, while most statistical comparisons indicated similar performance between testing environments for healthy and dementia participants, they were not all supported by strong evidence and certain comparisons even led to the opposite conclusion. Second, the present study was not ideally designed to compare the two testing environments, as the patient cohorts were different and the healthy control participants were not tested simultaneously in both environments within the same year. These findings would therefore need to be replicated in larger cohorts with the same patients in each test situation, to rule out the possibility of small differences observed in favour of face-to-face testing – and in particular, to assess the extent of individual variability in any differential effect of test environment.. Patients of equivalent disease severity would also need to be tested to compare the differential impact of diagnosis on remote performance over the course of the illness. Third, here we did not control for potential deficits in peripheral hearing as these are difficult to measure remotely without adequate equipment. Fourth, we manually adapted face-to-face tasks for remote administration, but there are now several established fully integrated online neuropsychological test batteries that have shown success in assessing patients with neurodegenerative disease remotely ^56^: future research could explore the extent to which our results are comparable with those obtained by such batteries.

Overall, the present findings demonstrate that, despite challenges in setting up remote testing protocols (specifically due to technological requirements), these produce similar results to face-to-face testing protocols. These are encouraging findings given the current climate and anticipating that research participants may continue to favour remote (or hybrid) visits over face-to-face assessments for reasons of convenience as well as safety, as we move beyond the COVID-19 pandemic.

## Data Availability

The data that support the findings of this study are available on request from the corresponding author.

## Acknowledgments

We are grateful to all participants for their involvement. The Dementia Research Centre is supported by Alzheimer’s Research UK, Brain Research UK, and The Wolfson Foundation. This work was supported by the Alzheimer’s Society, the Royal National Institute for Deaf People, Alzheimer’s Research UK, the National Institute for Health Research University College London Hospitals Biomedical Research Centre, and the University College London Leonard Wolfson Experimental Neurology Centre (grant PR/ylr/18575). MCRK is supported by a Wellcome Trust PhD Studentship (102129/B/13/Z). JJ is supported by a Frontotemporal Dementia Research Studentship in Memory of David Blechner (funded through The National Brain Appeal). EB was supported by a Brain Research UK PhD Studentship. RLB was supported by an MRC PhD Studentship in Mental Health. SJC was supported by grants from ESRC-NIHR (ES/L001810/1), EPSRC (EP/M006093/1) and Wellcome Trust (200783). CJDH was supported by a Royal National Institute for Deaf People–Dunhill Medical Trust Pauline Ashley Fellowship (grant PA23_Hardy) and a Wellcome Institutional Strategic Support Fund Award (204841/Z/16/Z).

## Competing interests

The authors have no competing interests to declare.

## Author contributions

MCRK, JJ, LD, EB, LR, RLB, EVB, SB, JDR, SJC, JDW and CJDH contributed to the conception and design of the study. MCRK, JJ, LD, EB, LR, RLB, EVB and CJDH contributed to the acquisition of data. MCRK, JJ, JDW and CJDH were involved in the analysis and interpretation of data, and wrote the first draft of the article. MCRK, JJ, LD, EB, LR, RLB, EVB, SB, JDR, SJC, JDW and CJDH were involved in revision of the draft and approved the final version of the manuscript.

## Notes

### Competing Interest Statement

The authors have declared no competing interest.

### Author Declarations

Ethical approval was granted by the University College London and National Hospital for Neurology and Neurosurgery Joint Research Ethics Committees in accordance with the Declaration of Helsinki.

