## Supplementary Material for "Neuropsychological assessments for dementia research in the COVID-19 era: comparing remote and face-to-face testing"

Table S1. List of general neuropsychological and neurolinguistic tests delivered face-to-face that were not included in the remote battery.

| **Test** | **Reason for removal** |
| --- | --- |
| WASI Vocabulary | Other tests from same domain already included |
| WASI Similarities | Other tests from same domain already included |
| Long Recognition Memory Test for Faces | To reduce the length of the remote battery, this was replaced with the Short Recognition Memory Test for Faces |
| Long Recognition Memory Test for Words | To reduce length of battery |
| Camden Paired Associates Learning | Other tests from same domain already included |
| WASI Block Design | Not feasible online as participants would not have blocks to manipulate |
| Stroop task | Not feasible online as too difficult for the examiner to determine the participant’s target and therefore whether or not they had made an error. Also differences in colour display across participants’ screens. |
| Trails A and B | Not feasible online as requires use pf pencil and paper |
| WAIS-R Digit Symbol | Not feasible online as requires use of pencil and paper |
| Usual/Unusual views | Other tests from same domain already included |
| Modified Kissing and Dancing | Other tests from same domain already included |
| Baxter Spelling Test | Not feasible online as face-to-face task requires the participant to use pencil and paper; typing responses was not considered appropriate due to potential interference from spell-check software |
| Written sentences | Not feasible online as face-to-face task requires the participant to use pencil and paper; typing responses was not considered appropriate due to potential interference from spell-check software |
| Spatial Span Forwards | To reduce length of battery |
| Spatial Span Backwards | To reduce length of battery |

We reduced the number of tests used in our face-to-face general neuropsychological and neurolinguistic batteries down for remote testing, reflecting a) the need to make the remote testing batteries shorter to minimise fatigue; b) impracticalities of administering certain stimuli remotely; and c) inability to adequately record participants’ responses to some tasks. The Table shows the tasks that were not included in the remote batteries.

Table S2. Performance on audibility screening task by remote participant groups

|  | CTL | AD | bvFTD | SD | PNFA | LPA |
| --- | --- | --- | --- | --- | --- | --- |
| Pre neuropsychology battery | | | | | | |
| Average number of incorrect items (/10) | 0.0  (0.0) | 0.1  (0.4) | 0.0  (0.0) | 0.0  (0.0) | 0.2  (0.4) | 0.0  (0.0) |
| Average number of errors on last three items (/3) | 0.0 (0.0) | 0.0 (0.0) | 0.0 (0.0) | 0.0 (0.0) | 0.0 (0.0) | 0.0 (0.0) |
| Pre neurolinguistic battery | | | | | | |
| Average number of incorrect items (/10) | 0.0  (0.0) | NA | NA | 0.0  (0.0) | 0.4  (0.9) | 0.0  (0.0) |
| Average number of errors on last three items (/3) | 0.0 (0.0) | 0.0 (0.0) | 0.0 (0.0) | 0.0 (0.0) | 0.0 (0.0) | 0.0 (0.0) |

The data indicate that there were no major background listening environmental confounds nor any significant differences between participant groups (all p>0.05). AD, patient group with typical Alzheimer’s disease; bvFTD, patient group with behavioural variant frontotemporal dementia; CTL, healthy control group; LPA, patient group with logopenic progressive aphasia; NA, not applicable; PNFA, patient group with progressive non-Fluent aphasia; SD, patient group with semantic dementia.

| **Test** | **CTL** | **All Patients** | **AD** | **bvFTD** | **SD** | **PNFA** | **LPA** |
| --- | --- | --- | --- | --- | --- | --- | --- |
| ***General intellect*** | | | | | | | |
| WASI Matrix | 2.56^t^ (Anecdotal) | 5.403 (Strong) | 3.134^t^ (Strong) | 1.477^t^ (Anecdotal) | 1.911^t^ (Anecdotal) | 1.356^t^ (Anecdotal) | 1.441^t^ (Anecdotal) |
| ***Episodic memory*** | | | | | | | |
| RMT Faces short | NA | 3.46 (Strong) | 0.411^t^ (Anecdotal) | NA | NA | NA | NA |
| ***Working memory*** | | | | | | | |
| DS (Forward) | 2.663^t^ (Anecdotal) | 4.54 ^t^ (Strong) | 3.467^t^ (Strong) | 1.948^t^ (Anecdotal) | 0.799^t^ (Anecdotal) | 2.528^t^ (Anecdotal) | 1.598^t^ (Anecdotal) |
| ***Language*** | | | | | | | |
| BPVS | 4.304^t^ (Strong) | 4.05 (Strong) | 3.363 (Strong) | 1.836 (Anecdotal) | 1.382^t^ (Anecdotal) | 2.754 (Anecdotal) | 1.261 (Anecdotal) |
| GNT | 2.923^t^ (Anecdotal) | 4.16 (Strong) | 3.542 (Strong) | 1.407^t^ (Anecdotal) | 2.331 (Anecdotal) | 1.68^t^ (Anecdotal) | 1.325^t^ (Anecdotal) |
| NART | 2.83^t^ (Anecdotal) | 1.99 (Anecdotal) | 1.552 (Anecdotal) | 1.511^t^ (Anecdotal) | 2.606^t^ (Anecdotal) | 2.52^t^ (Anecdotal) | 1.948^t^ (Anecdotal) |
| ***Arithmetic*** | | | | | | | |
| GDA Total | 3.671 (Strong) | 5.30 (Strong) | 3.33 (Strong) | 2.467^t^ (Anecdotal) | 0.943^t^ (Anecdotal) | 2.483^t^ (Anecdotal) | 1.123^t^ (Anecdotal) |
| ***Visuospatial*** | | | | | | | |
| VOSP | 0.0404 (Strong) | 4.76 (Strong) | 0.171^t^ (Strong) | 2.215 (Anecdotal) | 2.478 (Anecdotal) | 2.553 (Anecdotal) | 2.592^t^ (Anecdotal) |
| ***Executive*** | | | | | | | |
| DS (Reverse) | 4.304^t^ (Strong) | 5.28 (Strong) | 2.746^t^ (Anecdotal) | Variance at 0 | 1.991^t^ (Anecdotal) | 2.659 (Anecdotal) | 0.417^t^ (Anecdotal) |
| Letter fluency | 3.159^t^ (Strong) | 0.59 (Anecdotal) | 2.86^t^ (Anecdotal) | 2.426^t^ (Anecdotal) | 1.519^t^ (Anecdotal) | 0.511^t^ (Anecdotal) | 0.188^t^ (Strong) |
| Category fluency | 3.767^t^ (Strong) | 1.08 (Anecdotal) | 3.532 (Strong) | 2.186^t^ (Anecdotal) | 1.715^t^ (Anecdotal) | 1.027 (Anecdotal) | 1.29^t^ (Anecdotal) |

**Table S3. Bayesian statistics comparing general neuropsychological test performance on remote vs face-to-face assessments**

A Bayes factor (BF_01_) indicates the extent to which the null hypothesis is favoured against the alternative hypothesis (e.g., a BF_01_ value of 4 means that the obtained data are 4 times more likely under the null hypothesis than under the alternative hypothesis). A BF_01_ > 3 is therefore considered as strong evidence in support of the null hypothesis; while a BF_01_ of <1/3 is considered as strong evidence in support of the alternative hypothesis. Any values in between are categorised as ‘anecdotal’ evidence, equivalent to a non-significant result in inferential statistics ^1^. Results are influenced by the prior (more specifically the shape of the prior influences the strength of the evidence), which can be specified be default using a Cauchy distribution, as here; the Cauchy scale set here is 1.00. The superscript ^t^ indicates that a parametric Bayesian test was used; else the non-parametric Mann Whitney (with 1000 iterative samples) was employed. Blue shading indicates that the alternative hypothesis (H1, i.e. there was a difference in performance across the two environments) was favoured with strong evidence; Green shading indicates that the null hypothesis (H0; i.e. there was no difference in performance across environments) was favoured with strong evidence. AD, patient group with typical Alzheimer’s disease; BPVS, British Picture Vocabulary Scale; bvFTD, patient group with behavioural variant frontotemporal dementia; CTL, healthy control group; DS, Digit Span; F2F, face-to-face; GDA, Graded Difficulty Arithmetic test; GNT, Graded Naming Test; LPA, patient group with logopenic progressive aphasia; Matrix, WASI Matrix Reasoning; NART, National Adult Reading Test; PNFA, patient group with progressive nonfluent aphasia; RMT, Recognition Memory Test; SD, patient group with semantic dementia; VOSP, Visual Object Space Perception battery.

Table S4. Bayesian statistics comparing neurolinguistic test performance on remote vs face-to-face assessments

| **Test** | **CTL** | **All Patients** | **SD** | **PNFA** | **LPA** |
| --- | --- | --- | --- | --- | --- |
| ***Phoneme perception*** |  |  |  |  |  |
| PALPA 3 | 1.003 (Anecdotal) | 2.918 (Anecdotal) | 1.594 (Anecdotal) | 2.731 (Anecdotal) | 2.079 (Anecdotal) |
| ***Reading*** |  |  |  |  |  |
| Non word reading | 1.544 (Anecdotal) | 3.239 (Strong) | 1.940^t^ (Anecdotal) | 2.033^t^ (Anecdotal) | 2.428^t^ (Anecdotal) |
| Regular reading | Variance at 0 | 2.989 (Anecdotal) | 1.975 (Anecdotal) | 2.305 (Anecdotal) | 1.923 (Anecdotal) |
| Irregular reading | Variance at 0 | 2.996 (Anecdotal) | 2.256 (Anecdotal) | 2.194^t^ (Anecdotal) | 2.005 (Anecdotal) |
| ***Naming*** |  |  |  |  |  |
| BNT | 3.612^t^ (Strong) | 1.995 (Anecdotal) | 0.584^t^ (Anecdotal) | 2.580 (Anecdotal) | 1.490 (Anecdotal) |
| ***Semantic association*** |  |  |  |  |  |
| Camel and cactus | 4.039^t^ (Strong) | 1.067^t^ (Anecdotal) | 1.524^t^ (Anecdotal) | 2.102 (Anecdotal) | N<2 for F2F |
| ***Word comprehension*** |  |  |  |  |  |
| Concrete synonyms | 2.739^t^ (Anecdotal) | 3.622 (Strong) | 2.009^t^ (Anecdotal) | 2.400^t^ (Anecdotal) | 1.413^t^ (Anecdotal) |
| Abstract synonyms | 1.726^t^ (Anecdotal) | 2.718 (Anecdotal) | 0.782 (Anecdotal) | 2.741 (Anecdotal) | 1. 941 (Anecdotal) |
| ***Sentence comprehension*** |  |  |  |  |  |
| PALPA55 | 0.944 (Anecdotal) | 3.954 (Strong) | 2.246 (Anecdotal) | 2.603 (Anecdotal) | 2.110 (Anecdotal) |
| ***Speech repetition*** |  |  |  |  |  |
| Monosyllabic word repetition | 0.0487^t^ (Strong) | 0.117 (Anecdotal) | 0.477^t^ (Anecdotal) | 1.167^t^ (Anecdotal) | 1.055^t^ (Anecdotal) |
| Bisyllabic word repetition | 4.304^t^ (Strong) | 3.744 (Strong) | 1.086 (Anecdotal) | 2.639 (Anecdotal) | 0.650^t^ (Anecdotal) |
| Trisyllabic word repetition | Variance at 0 | 3.701 (Strong) | 1.339 (Anecdotal) | 2.400 (Anecdotal) | 1.677 (Anecdotal) |
| Graded difficulty sentence repetition | 1.779 (Anecdotal) | 2.983 (Anecdotal) | 1.420^t^ (Anecdotal) | 2.535^t^ (Anecdotal) | 1.996 (Anecdotal) |
| ***Sentence construction*** |  |  |  |  |  |
| Spoken | Variance at 0 | 1.525 (Anecdotal) | 2.270 (Anecdotal) | 1.960. (Anecdotal) | 0.930^t^ (Anecdotal) |

A Bayes factor (BF_01_) is shown for each remote vs face-to-face testing comparison. The interpretation and colour coding are as indicated in the legend to Table S3 above. BNT, Boston Naming Test; CTRL, healthy control group; F2F, face-to-face; LPA, patient group with logopenic progressive aphasia; N, number of participants per group; PALPA, Psycholinguistic Assessment of Language Processing in Aphasia subtests; PNFA, patient group with progressive nonfluent aphasia; SD, patient group with semantic dementia

Figure S1. Example of basic audibility screening measure

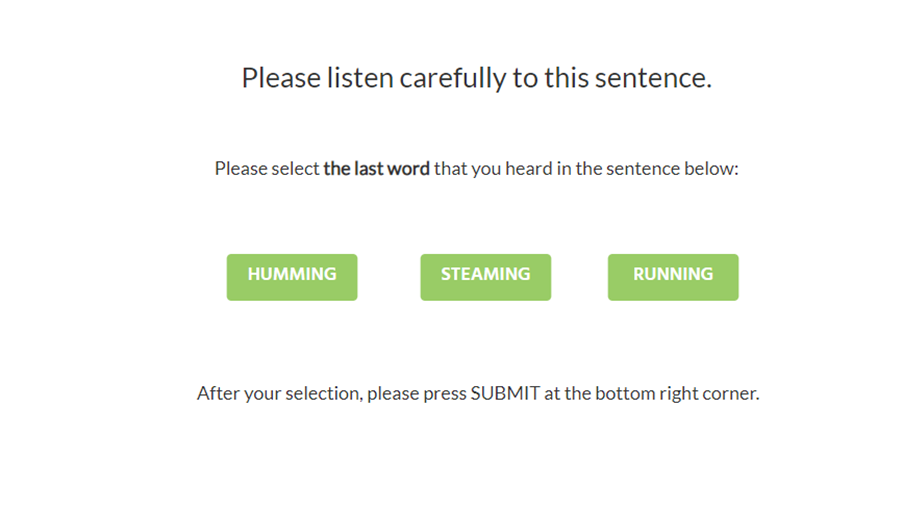

The Figure shows a Labvanced display of the BKB hearing screening measure. In this example, the sentence spoken was “The car engine is running”. For each sentence, two foils were displayed alonside the target, both of which made sense in the sentence when replacing the target. One of the foils was also selected to loosely rhyme with the target word (here, “humming”).
